# Estimating Burden of Mortality due to Excess Body Mass Index in the US Adult Population by Combining Evidence from a Mendelian Randomization Study and National Health Surveys

**DOI:** 10.1101/2023.03.17.23287394

**Authors:** Prosenjit Kundu, Stephen Burgess, Nilanjan Chatterjee

## Abstract

**Importance:** Assessment of the burden of mortality due to excess body weight in a population and its subgroups is important for designing health policies for interventions. Mendelian randomization (MR) studies can provide an opportunity to correct for unmeasured confounding bias present in observational studies, but such evidence has not been used to assess population burden of mortality due to excess BMI.

**Objective:** Combine results from a recent Mendelian randomization (MR) study and data from the National Health Surveys to estimate preventable fraction (PF) of 10-year all-cause and cause-specific mortality by different degrees of BMI reduction in the US adult population and underlying risk strata.

**Designs:** We use cross-sectional data on the distribution of BMI and other risk factors of mortality from the National Health and Nutritional Examination Surveys (NHANES) across two-time spans (1999-2006 and 2017-2018). We use linked data from National Death Index to characterize the observed risk of 10-year mortality associated with BMI and other risk factors based on the NHANES 1999-2006 cohort. We further import results from an external MR study on linear and non-linear effects of BMI and use novel methods to estimate preventable fraction (PF) for deaths under different counterfactual scenarios of BMI reduction in the NHANES population.

**Settings:** Primary analysis is restricted to the NHANES non-Hispanic white population (age range 40-69 years) due to the unavailability of MR studies in other groups, but projections are provided for the African American population under the assumption of homogeneity of causal effects.

**Outcome:** Preventable fraction for 10-year all-cause mortality and cause-specific mortality due to 50% and 100% reduction of excess BMI (BMI>25.6 kg/m^2^) for the US adult population in the age range of 40-69 years.

**Results:** Nearly 33% and 43% of the NHANES 2017-2018 target population are overweight (25.6 kg/m^2^ ≤BMI<30.7 kg/m^2^) and obese (BMI>30.7 kg/m^2^), respectively, according to WHO definitions. Estimates of relative risks for different BMI categories (relative to normal BMI) from the external MR study range from 1.05 (25.6 kg/m^2^ ≤ BMI < 27.8 kg/m^2^) to 5.95 (BMI> 42.4 kg/m^2^). We estimate PF for 10-year all-cause mortality due to 50% and 100% reduction of excess BMI for the population to be 24% (95% CI: 14 – 34) and 35% (95% CI: 22−48), respectively. The estimate of PF of death due to heart disease and cancer for this population reaches up to 48% (95% CI: 25≤71) and 18% (95% CI: -2−38), respectively. Partitioning of PF shows that 60% of all BMI-attributable deaths arise from only 12% of the population who are at the highest risk due to obesity and a combination of other risk factors.

**Conclusions:** Nearly one in three deaths in a contemporary US adult population can be attributed to overweight and obesity. A substantial fraction of these deaths are likely to be preventable through pragmatic and targeted BMI interventions.

## Introduction

Body mass index, defined as weight measured in kilograms as a ratio to squared heights measured in meters, has been widely associated with the risk of a variety of chronic diseases and subsequent prognosis^1-4^, as well as serious illness associated with the ongoing pandemic of COVID-19^5-7^. Recent large international studies have shown that the prevalence of overweight and obesity are increasing worldwide, and associated health effects have been described as a global pandemic^8^. As BMI affects a wide variety of health outcomes, estimation of its impact on total mortality has been considered critical to understanding the total magnitude of associated health burden^9,10^.

A variety of studies have previously attempted to estimate the number of deaths attributable to overweight, obesity, and sometime under-weight at national and international levels, but results across studies have often been inconsistent due to methodological issues^9-18^. In the US alone, for example, estimates of preventable fraction (PF), also known as population attributable fraction (PAF) and population attributable risk (PAR), of death due to obesity have been reported to range between 5-20% across studies^19^. A major source of heterogeneity is the underlying estimates of risks associated with different BMI categories, which are typically obtained from observational epidemiologic studies and hence subject to confounding and reverse causality bias. More recent studies have attempted to remedy the problem by the estimation of BMI risk parameters by pooling data from a large number of cohort studies and restricting the analysis to healthy individuals^9,12^, but the concern of bias due to confounding remains. A second issue has been the use of an incorrect formula for the calculation of PF when using estimates of BMI risk parameters that have been adjusted for known confounders^19^.Recently a series of Mendelian Randomization (MR) studies have reported estimates of the effect of genetically predicted BMI on the risk of mortality and a wide variety of health outcomes^20-26^. As genetically predicted BMI is immune to reverse causality and less likely to be influenced by unmeasured confounding, the reported estimates provide an opportunity to re-evaluate deaths attributable to BMI potentially correcting for biases persistent in purely observational studies. Further, prior studies have exclusively reported on PAF/PF that corresponds to many potential deaths preventable by the reduction of BMI of all overweight and obese individuals to the lowest-risk level. For understanding the potential impact of pragmatic interventions, however, there is an imminent need for quantifying the burden of deaths preventable through a more modest degree of BMI reduction, taking into account the nature of the full spectrum of a dose-response relationship in underlying risk.

In this article, we develop and apply novel methods to derive estimates of preventable fraction (PF) of deaths by different levels of BMI reduction in the US adult population by combining evidence from a recent MR study of linear and non-linear effects of BMI on mortality, and individual level data on BMI, co-factors and linked mortality outcome data from the National Nutritional and Health Examination Survey (NHANES). To explore opportunities for targeted interventions, we further provide partitioning of PF by risk strata defined by a combination of sociodemographic and lifestyle factors and estimates of absolute risk reduction achievable by different degrees of BMI reduction across these strata. These analyses provide new insights into the population- and individual-level impact of potential interventions for BMI reduction. The methodologic framework we develop can be applied broadly to perform similar calculations for other health outcomes and modifiable risk factors.

## Methods

### Data Sources

*MR Estimates for BMI-effects on Mortality*. Sun et al^25^ recently reported estimates of the effects of WHO-defined BMI categories on the risk of mortality based on linear and non-linear MR analysis conducted across two cohort studies: HUNT and UK Biobank. The MR estimates, which represent the effect associated with genetically predicted BMI, are expected to represent the causal effect of a lifelong change in BMI^27^. We use results from the analysis of UKBiobank which have more similar population characteristics as that of the US target population of interest. The study participants in UKBiobank are middle to early late-aged individuals (40-69 years) of largely European ancestry (∼ 95%) and include a total number of deaths of 10,344 accrued over a median follow-up of 7 years. In both studies, the non-linear MR analysis identified a similar J-shaped relationship between risk of mortality in the overall population, though the hazard-ratio estimates for obese (BMI> 30) categories appeared to be stronger in the UK Biobank study (see Figure 2 in Sun et al.^25^). Further, in subgroup analysis, the J-shaped relationship was only found to be present among smoker, but an ever-increasing relationship appeared among neve smokers. We obtained reported MR estimates of BMI-associated hazard ratios for all-cause mortality and two cause-specific mortality (heart disease and cancer) and all corresponding estimates of variance-covariance terms in the subsequent derivation of PF estimates and their standard errors.

### National Health and Nutritional Examination Surveys (NHANES)

NHANES is a cross-sectional survey which biennially collects information on health-related factors using complex sampling designs. The sample is representative of the noninstitutionalized U.S. civilian population residing in the 50 states and the District of Columbia^28,29^. National Center of Health Statistics (NCHS) links the survey data to the National Death Index (NDI) database to efficiently collect information on mortality^30^. We use the cross-sectional data available from the NHANES across two-time frames, 1999-2006 and 2017-18 to estimate the prevalence of individuals in different BMI categories, excluding underweight individuals (≤ 22.06 kgm^-2^).

We use follow-up data on 10-year mortality for the 1999-2006 cohort, available through linkage to the 2015 National Death Index database^30^ to model and estimate observed risk (potentially confounded) of 10-year all-cause and cause-specific mortality associated with the BMI categories for the NHANES population – a key ingredient needed for valid PF calculation (see **Methods**). We assume the observed risk of 10-year mortality associated with BMI is the same for the 1999-2006 and 2017-18 cohorts, as sufficient follow-up data are not available for the later cohort. We restricted the analysis to individuals in the age range of 40-69, same as that of the UK Biobank study from which the MR estimates of BMI effects were derived. We also used data from the 1999-2006 cohort to estimate the multi-factorial risk of 10-year all-cause and cause-specific mortality associated with a number of other key risk factors of mortality, including age, gender, smoking, alcohol consumption, education, marital status, and use an underlying risk-score to define population strata. All modeling is performed using logistic regression and it is assumed that underlying odd-ratios provide good approximations to the risk ratios as the overall 10-year all-cause mortality rate for the population over ten years is low (11%). As MR estimates for BMI effects were derived from studies consisting mostly of European ancestry individuals, we first estimated the PF based on the NHANES non-Hispanic white participants only. In a secondary analysis, we projected PF for the NHANES non-Hispanic black population assuming that the available MR estimates for BMI effects can also apply to this population, but the underlying assumption needs to be tested in the future.

### Statistical Methods

Traditionally, *Preventable fraction (PF)* for a binary outcome (*Y*), e.g death, associated with an exposure (*X*) in reference to an exposure value of *X* = *x*_0_ corresponding to the lowest risk is defined as the proportion of the outcomes that would be prevented if the exposure value of all individuals in the population were set to *X* = *x*_0_. Mathematically, the definition corresponds to

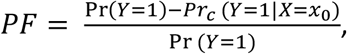

*Pr*_*c*_ (*Y* = 1|*X* = *x*_0_) is the probability of the outcome in a “counterfactual” population where the exposure values for all individuals have been set to *X* = *x*_0_. In the presence of other co-factors, such as confounders (measured or unmeasured) and effect modifiers, the *PF* quantifies the proportion of the adverse outcomes that will be reduced in a hypothetical intervention that allows to shift the distribution of the exposure in the population to the lowest value *X* = *x*_0_, *without affecting the distribution of the other factors*. In general, Pr (*Y* = 1|*X* = *x*_0_), the observed risk of the outcome for individuals with *X* = *x*_0_, is not the same as *Pr*_*c*_(*Y* = 1|*X* = *x*_0_).

In the absence of other co-factors, Levin (for binary exposure)^31,32^ and Walter (for multiple categories of exposure)^18,33^ showed that the PF can be conveniently represented in terms of relative risk of the outcome associated with the exposure and the exposure prevalence in the underlying population (**see formula (2) in Supplemental Methods**). In the presence of confounders, however, it is known that this formula does not represent PF even if the relative-risk parameters were to be adjusted for all potential confounders^18^. Yet the formula continues to be used widely, including in the recent WHO global burden of disease study^9^, as it allows convenient combining of estimates of exposure prevalence and relative risk parameters from separate epidemiologic studies.

In this article, we derive a valid representation of PF (**see formula (5) in Supplemental Methods**) in terms of (i) exposure prevalence for the underlying target population (e.g. NHANES) (ii) unadjusted/raw relative-risk parameters associated with the exposure, also in the underlying target population (NHANES) and (iii) additional estimates of relative-risk parameters from external studies, such as a randomized trial or an MR study, that can be considered to be representing the unconfounded effect of the exposure and is transportable to the target population. We show that the formula can also be alternatively derived from another relatively unknown representation of PF^34^ (also called Bruzzi’s formula, see section 2 (b) in Supplemental Methods) that allows valid estimation of it in the presence of confounders. This representation requires estimates of exposure prevalence among cases (Y=1) in the underlying target population in addition to those for the general population and confounding adjusted relative-risk parameters. The proposed formula (5), however, is more flexible and allows the use of models in estimating observed risks.

We further use the framework to derive formulae for individual-level absolute risk reduction due to lowering of BMI by a given amount, population-level PF associated with any downward shift of BMI distribution, and partitioning of PF according to risk-strata (**see formula 7-9 in Supplemental Methods**). All confidence intervals are derived based on the delta method^35^ accounting for sources of uncertainty associated with both the NHANES study and the external MR estimates (see Section 3 in Supplemental Methods). All the estimates of PF are shown in terms of percentage unless otherwise stated.

## Results

Characteristics of the NHANES white study participants from 1999-2006 who have normal or higher BMI are shown in **Supplementary Table 1**. Overall BMI appears to be associated with a number of risk factors including gender, smoking behavior, alcohol intake, education, and marital status. Relative risks of mortality associated with BMI categories are shown in **Table 1** based on three different sources (1) internal analysis of NHANES data (2) a large external pooled cohort analysis^9^ and (3) external MR analysis (Table 1). Overall, relative risks or excess BMI categories are notably stronger from the MR analysis than that from the internal NHANES analysis for obese categories (≥ 30.73 Kgm^-2^). Estimates from pooled cohort analyses are close to those from the MR analysis for overweight categories but are attenuated for the obese categories. The relative-risk estimates shown in **Table 1** from the NHANES and external MR analyses, and the corresponding prevalence of BMI categories form the basic input data for our main analysis.

**Table 1:** Population prevalence and risk of mortality (95% CI) associated with BMI categories. Prevalence of BMI categories are derived from the NHANES 1999-2006/2017-18 data. Relative risks of mortality for BMI categories are calculated with respect to the reference category 22.06 – 25.66 Kgm-2. Three sets of relative-risk parameters correspond to (1) internal analysis of NHANES 1999-2006 data linked to mortality outcomes; (2) results reported from a large external pooled cohort study and (3) external Mendelian randomization study.

| BMI categories |  | 25.66 – 27.78 | 27.78 – 30.73 | 30.73 – 36.13 | 36.13 – 42.35 | ≥ 42.35 |
| --- | --- | --- | --- | --- | --- | --- |
| Prevalence of the BMI categories in NHANES<sup>1</sup> 1999-2006/2017-18(in %) |  | 17/12 | 21/21 | 23/26 | 10/10 | 4/7 |
|  |  | Odds ratio |  |  |  |  |
| All-cause mortality | NHANES<sup>1</sup> (95% CI) | 1.25<br>(0.81-1.93) | 1.25<br>(0.90 – 1.73) | 1.32<br>(0.94 – 1.88) | 2.15<br>(1.56-2.95) | 2.20<br>(1.30 – 3.72) |
|  | Pooled<sup>2</sup> estimate (95% CI) | 1.06<br>(1.05 - 1.07) | 1.19<br>(1.17 – 1.22) | 1.45<br>(1.41 – 1.50) | 1.96<br>(1.86 – 2.06) | 2.89<br>(2.72 – 3.07) |
|  | MR estimate (95% CI) | 1.05<br>(0.99 – 1.11) | 1.20<br>(1.05 – 1.36) | 1.74<br>(1.26 – 2.41) | 3.49<br>(1.83 – 6.66) | 5.95<br>(2.57 – 13.78) |
| Heart-disease mortality | NHANES<sup>1</sup> (95% CI) | 1.55<br>(0.58 – 4.17) | 1.96<br>(.84 – 4.57) | 1.80<br>(0.72 – 4.51) | 4.66<br>(2.24 – 9.69) | 4.84<br>(1.66 – 14.01) |
|  | Pooled<sup>2</sup> estimate (95% CI) | 1.12<br>(1.11 – 1.13) | 1.34<br>(1.30 – 1.38) | 1.72<br>(1.65 – 1.79) | 2.67<br>(2.48 – 2.88) | 3.71<br>(4.05 – 4.41) |
|  | MR estimate (95% CI) | 1.09<br>(1.04 – 1.17) | 1.30<br>(1.02 – 1.65) | 2.00<br>(1.07 – 3.74) | 4.44<br>(1.17 – 16.84) | 8.21<br>(1.34 – 50.27) |
| Cancer mortality | NHANES<sup>1</sup> (95% CI) | 1.29<br>(0.72 – 2.34) | 1.14<br>(0.72 – 1.82) | 0.97<br>(0.61 – 1.54) | 1.49<br>(0.80 – 2.78) | 1.02<br>(0.35 – 3.00) |
|  | Pooled<sup>2</sup> estimate (95% CI) | 1.04<br>(1.01 - 1.06) | 1.12<br>(1.10 - 1.15) | 1.28<br>(1.24 - 1.33) | 1.56<br>(1.48 – 1.64) | 1.86<br>(1.73 – 2.00) |
|  | MR estimate (95% CI) | 1.03<br>(0.95 – 1.11) | 1.11<br>(0.93 – 1.32) | 1.32<br>(0.90 – 1.94) | 1.78<br>(0.86 – 3.68) | 2.26<br>(0.89 – 5.77) |

<sup>1</sup> All analyses of NHANES data incorporated sample weights. The odds ratios from the NHANES analysis are not adjusted for any other factors and these “raw” estimates are incorporated into PF calculations according to formula (5) of Supplemental Methods.
<sup>2</sup>Results are based on analysis of all Western cohorts as reported in Di Angelantonio, E., et.al<sup>9</sup>. The BMI categories used in this study are similar to the ones used for the external MR study, but they are not identical. Note that the pooled estimates are based on analysis in never-smokers without known chronic disease at baseline and excluding the first 5 years of follow-up.

Our estimate of PF_100%_, which corresponds to the usual definition of PF/PAR, indicates that more than one in four deaths could be attributed to excess BMI for the self-reported non-Hispanic white population in the age range of 40-69 represented by the NHANES 1999-2006 surveys (**Table 2**). Further, these estimates imply that for the same population, every one in three and one in six deaths due to heart disease and cancer, respectively, can be attributed to excess BMI. When accounting for the increasing prevalence of overweight and obesity in the more recent (2017-18) population, the PF for excess BMI increases notably for all outcomes and populations (smokers and non-smokers). Overall, our estimates imply that every one in three future deaths for the current US population could be implicated to excess BMI.

**Table 2:** Estimates of Preventable Fraction (PF) for 10-year mortality in self-reported non-Hispanic White individuals and age group 40-69 in the NHANES populations for the time periods 1999-2006 and 2017-18. Two sets of estimates of PF (in %) are obtained corresponding to 100% (PF_100%_)^1^ and 50% (PF_50%_)^2^ reduction of excess BMI (compared to normal weight) across all individuals in the underlying populations. Results are derived using BMI prevalence data from NHANES, linked mortality outcome data for the 1999-2006 NHANES cohort, and estimates of BMI effects from an external MR study. The reference categories for BMI are chosen according to those provided by the MR study: 22.06 – 25.66 Kgm^-2^ for the whole population, 22.3 – 26 Kgm^-2^ for ever-smokers, and 21.9 – 25.4 Kgm^-2^ for never-smokers. All analyses of NHANES data incorporate sampling weights.

| Mortality outcome | Smoking status | Estimate (in %) of $PF_{100\%}$ (95% CI) | | Estimate (in %) of $PF_{50\%}$ (95% CI) | |
| --- | --- | --- | --- | --- | --- |
|  |  | 1999-2006 | Projected for 2017-18 | 1999-2006 | Projected for 2017-18 |
| All-cause | All | 30<br>(17 – 44) | 35<br>(22 – 48) | 21<br>(9 – 33) | 24<br>(14 – 34) |
|  | Ever <sup>a</sup> | 28<br>(12 – 44) | 34<br>(18 – 50) | 20<br>(4 – 37) | 24<br>(12 – 36) |
|  | Never | 25<br>(-7 – 57) | 32<br>(-2 – 65) | 16<br>(-10 – 41) | 16<br>(-12 – 43) |
| Heart disease | All | 42<br>(20 – 64) | 48<br>(25 – 71) | 30<br>(8 – 52) | 32<br>(9 – 54) |
| Cancer | All | 17<br>(-2 – 36) | 18<br>(-2 – 38) | 10<br>(-3 – 23) | 11<br>(-2 – 24) |

<sup>1</sup> $PF_{100\%}$ for 1999-2006 and for 2017-18 are respectively calculated using the formula (5) and formula (17) provided in the supplementary methods. See Section 4 of Supplemental Methods for details.
<sup>2</sup> $PF_{50\%}$ for 1999-2006 using formula (9) of supplementary methods. $PF_{50\%}$ projected for 2017-18 is calculated using formula (18) provided in the supplementary methods. See Section 4 of Supplemental Methods for details.
<sup>a</sup> Ever smokers include former and current smokers.

We inspect PF_50%_ as a measure of preventable deaths achievable possibly through a more pragmatic intervention that leads to a 50% reduction of excess BMI across the board. Encouragingly, our estimates indicate that in a contemporary population, close to 70% ((24/35)× 100) of BMI-attributable deaths could potentially be prevented by only a 50% reduction of excess BMI. We further partition total preventable deaths due to excess BMI by the combination of overweight categories and quintiles of a risk score that is defined by other risk factors of mortality, assuming that MR-estimated BMI relative-risk parameters are applicable across risk strata (**Table 3**). Results demonstrate a large fraction of preventable deaths due to excess BMI could arise from a relatively small fraction of the highest-risk population. For example, individuals who are obese and at the highest quintile of the risk score, represent only 6% of the population but are expected to give rise to 42% of BMI-attributable deaths.

**Table 3:** Partitioning of all-cause deaths attributable to excess BMI by overweight categories and risk-score quintiles defined by other prominent risk factors of mortality. The analysis is based on NHANES 1999-2006 cohort with a reference category for BMI of 22.06 – 25.66 Kgm^-2^. All analyses of NHANES data incorporate sampling weights.

| Quintiles of risk score defined by other risk-factors<sup>1</sup> | BMI category |  |
| --- | --- | --- |
|  | Pre-obese<sup>2</sup> (25.66-30.72 Kgm<sup>-2</sup>)<br>% Of Population/<br>% Of BMI-attributable deaths | Obese<sup>2</sup> (>=30.72 Kgm<sup>-2</sup>)<br>% Of Population/<br>% Of BMI-attributable deaths |
| **1** | 8.86/0.55 | 8.78/3.80 |
| **2** | 8.72/1.46 | 8.81/10.69 |
| **3** | 8.14/2.16 | 7.37/14.89 |
| **4** | 6.94/2.51 | 6.57/19.43 |
| **5** | 5.74/2.96 | 5.90/41.55 |

<sup>1</sup> Other risk factors include age, sex, smoking status, average number of cigarettes smoked per day, alcohol status, education status, and marital status.
<sup>2</sup> The categories from the external MR study based on their BMI percentiles are collapsed and closely correspond to WHO (https://www.who.int/europe/news-room/fact-sheets/item/a-healthy-lifestyle---WHO-recommendations) defined categories which are 25-30 Kgm<sup>-2</sup> for pre-obese and >=30 Kgm<sup>-2</sup> for Obese (including Obese grade I, grade II and grade III).

Estimates of absolute risks show a potential level of 10-year all-cause and cause-specific mortality risk reductions achievable at an individual level by lowering BMI among obese individuals (**Figure 1**, see **Supplementary Figures 1-2** for cause-specific mortalities). For the non-Hispanic white population in the age range of 40-69 represented by the NHANES 1999-2006 surveys, we estimate an average absolute risk reduction for the 10-year all-cause mortality of 0.04 and 0.06 corresponding to excess BMI reduction of 50% and 100%, respectively (**Figure 1**). The level of BMI-induced risk reduction, however, is expected to vary widely according to underlying risk defined with other causes of mortality. Individuals at the top 20% or higher risk of 10-year mortality due to causes other than BMI, for example, are expected to observe a lowering of absolute risk by as much as 0.11 and 0.14 corresponding to excess BMI reduction of 50% and 100%, respectively. In contrast, individuals at the bottom 20% of risk due to other causes, will only expect absolute risk reduction of 0.009 and 0.012 for the same degrees of BMI reduction, respectively.

**Figure 1:**
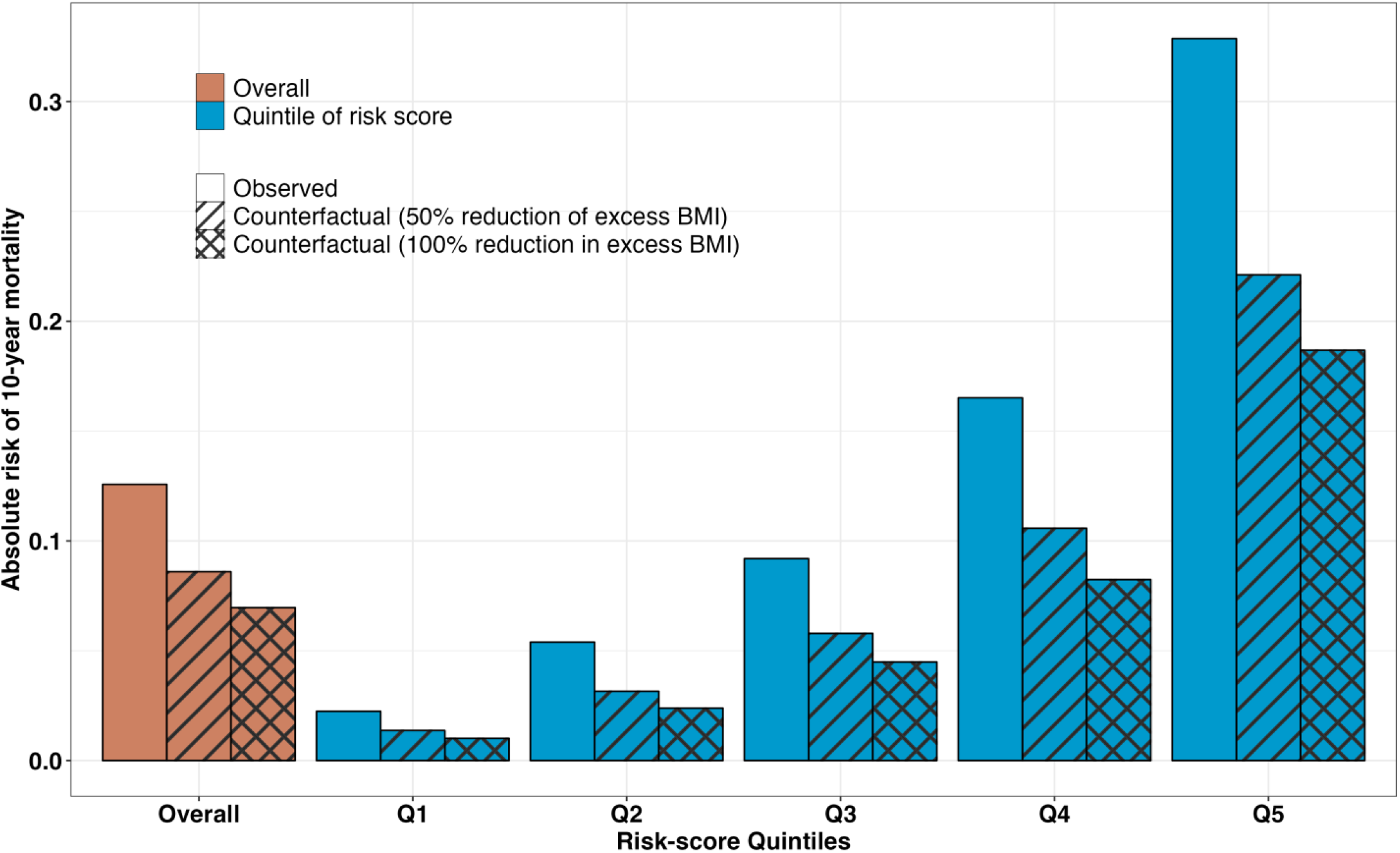
Observed and counterfactual absolute risks of 10-year mortality for obese individuals in the non-Hispanic White population with an age range of 40-69 represented by the NHANES 1999-2006 Surveys. Observed risks correspond to empirical proportions of deaths observed. Counterfactual risks correspond to the average of the estimated absolute risks of the individuals associated with 50% and 100% reduction of excess BMI (compared to normal weight) where MR-derived estimates of relative risks are used to represent underlying counterfactual effects. Results are shown for the overall population and stratified by the quintile of a risk score defined by other prominent risk factors of mortality. See Sections 1 and 4 of Supplemental Methods for details of the derivations.

As external MR studies have been primarily conducted based on European ancestry populations, we have restricted our main analysis to the NHANES non-Hispanic white population only. We obtained extrapolated estimates of PF for all-cause mortality for the NHANES non-Hispanic black population assuming that the MR estimates for relative risks for different BMI categories could be also applicable to this population (See **Supplementary Table 2-3** for population characteristics and required input data). As expected, due to the higher prevalence of obesity, the estimates of PF for the non-Hispanic black population are higher compared to those for the white populations during 1999-2006 (**Supplemental Table 4**). However, for the more recent NHANES population (2017-2018), the projected PFs are similar for the two populations.

Finally, in a sensitivity analysis, we obtain estimates of PF and individualized absolute risks based on estimates of relative risks from BMI categories obtained from a large, pooled cohort analysis^9^ (see **Table 1** and **Supplementary Table 5**). We use the same methodology as the main analysis, except that the input for “fully adjusted” relative risks is derived from the pooled cohort studies instead of MR analysis. We observe that estimates of PF (**Supplemental Table 5**) and degree of absolute risk reductions (**Supplemental Figure 3-5**) decrease by a notable extent compared to those derived based on MR estimates of relative risks (**Table 2, Figure 1, Supplemental Figure 2-3**). In particular, for the contemporary population of 2017-18, we observe estimates of PF decrease by 26%, 15%, and 11% for all-cause, heart-disease and cancer deaths, respectively, due to the use of estimates of relative risks from the pooled cohort studies compared to those from the MR study.

## Discussion

In this study, we evaluate the burden of mortality due to excess BMI in the US population, and opportunities for prevention, through a series of novel analyses which combine data from nationally representative health surveys and evidence from an external Mendelian-Randomization study. Our estimates of PF indicate that about 1 in 3 future deaths in a contemporary US population can be attributed to excess BMI, and yet, encouragingly, nearly 70% of the BMI-attributable deaths could be prevented by only a 50% reduction of excess BMI for the population. Further, the partitioning of deaths attributable to excess BMI by other risk factors of mortality shows that a large fraction of BMI-attributable deaths is expected to arise from a small fraction of the population, indicating opportunities for targeted interventions. Finally, estimates of absolute risks in various counterfactual settings show that the degree of risk reduction achievable at an individual level by lowering BMI is expected to vary widely by the risk associated with other factors.

As indicated earlier, previous estimates of PF of mortality due to BMI in the US and other populations have varied widely due to a number of methodological issues. The most significant challenge has been dealing with bias associated with the estimation of BMI effects on mortality due to confounding and reverse causality in observational studies. Generally, studies that restricted the analysis to non-smokers and healthy individuals produced larger estimates of relative risks and PF^9,36,37^. A very large international pooled cohort analysis study^9^, for example, used such restrictions to report relative risks of mortality for different BMI categories (see **Table 1**). These estimates of relative risks, which are attenuated compared to those from the MR study for the highest obese groups, result in a considerably lower estimate of PF in our analysis, but not beyond limits of uncertainty (**Table 2** and **Supplemental Table 5**).

Direct comparison of PF estimates from our study with others, however, is challenging for a number of reasons. MR estimates correspond to the effects of genetically predicted BMI which remains stable over the time course, our results should be interpreted as PF due to life-long changes in BMI^27^. In epidemiologic cohort studies, on the other, BMI measurements typically correspond to values observed at a single time point and thus may not capture the true effect of long-term exposure. Further, many external studies continue to use an incorrect formula for the evaluation of PF when relative-risk estimates are adjusted for potential confounders^19^. Estimates of PF from different formulae within our application clearly show that the use of the incorrect formula can lead to a very substantial upward bias (**Supplementary Table 6**).

The traditional definition of PF, also PAR and PAF, provides a measure of the total burden of deaths that could be preventable by shifting the BMI of an entire population to normal weight. But such measures do not provide an assessment of opportunities for prevention through pragmatic interventions that are likely to lead to a more modest reduction in BMI. Our estimates of PF_50%_ show that a majority (70 %) of the BMI-attributable deaths could be prevented by lowering excess BMI by 50% -a more realistic target for a population based on existing interventions. At an individual level, we observe that our estimate of risk reduction of mortality by 50% lowering of excess BMI among obese individuals, which roughly corresponds to a 17% reduction of net BMI (**Figure 1**), is fairly consistent with the range of estimates of long-term benefit from Bariatric surgery reported from randomized trial^38-40^ and well controlled observational studies^40,41^. A similar reduction in body weight is seen with anti-obesity medication, Semaglutide, reported from a randomized clinical trial, however, future research is merited to look at the long-term survival benefits attributed to body weight reduction from such medication. ^42^. Further, we show that the net benefit from such interventions is expected to vary widely across individuals according to their risks associated with other factors. Overall, these results suggest that it should be possible to save a large number of BMI-attributable deaths in the population through a realistic reduction of BMI and there are considerable opportunities for targeted interventions.

Our study has several limitations. First, the total sample size and the number of observed deaths for the NHANES 1999-2006 study population, which influences the estimation of both prevalence of BMI categories and underlying observed relative-risk parameters, are moderate and thus lead to wide confidence intervals in some of the analyses (e.g., cancer-specific deaths). Second, in partitioning/stratifying the burden of BMI by other risk factors, we implicitly assume an underlying multiplicative model and that the external MR estimates of relative risk for BMI categories can be applied across the risk categories. Future MR studies of BMI are needed for more in-depth investigation of the potential heterogeneity of underlying causal effects of BMI by other major risk factors. Similarly, in projecting the burden of deaths for the non-Hispanic black population, we had to assume the applicability of the MR estimates of BMI effects, which were derived mostly from European ancestry studies. As GWAS continues to grow in African and other non-European ancestry populations, it will be imperative to investigate the potential for BMI by ancestry interactions on the risk of 10-year mortality through MR analyses.

In conclusion, we carry out a series of novel analyses to examine the population- and individual-level impact of excess BMI in the US adult population by combining data from National Health Surveys and recent Mendelian Randomization studies. These analyses, while shows that the population faces a very large burden of mortality due to excess BMI, there are promising opportunities to save many lives through realistic BMI reduction and targeted interventions. Our analytic framework could be used in the future to assess the challenges and opportunities for the prevention of a wide variety of health outcomes associated with modifiable risk factors.

## Data Availability

All data produced in the present work are contained in the manuscript.

https://pubmed.ncbi.nlm.nih.gov/30957776/

## Acknowledgement

This work was funded by NIH grants: R01 HG010480-01 (NC) and U01 HG011719 (NC and PK).

## Author Contributions

PK and NC had full access to all of the data in the study and take responsibility for the integrity of the data and the accuracy of the data analysis.

*Concept and design:* PK and NC

*Acquisition, analysis, or interpretation of data*: PK, SB and NC

*Drafting of the manuscript:* PK and NC

*Critical revision of the manuscript for important intellectual content:* PK, SB and NC

*Statistical analysis*: PK

*Obtained funding*: NC

*Administrative, technical, or material support*: PK, SB and NC

*Supervision*: NC

