## Supplemental Methods for "Estimating Burden of Mortality due to Excess Body Mass Index in the US Adult Population by Combining Evidence from a Mendelian Randomization Study and National Health Surveys"

### 1. Key formulae

In this section, we describe the key formulae used in the manuscript for calculating total PF, partitioning of PF by risk groups, and individual-level risk differences. We derive the formulae assuming a categorical exposure $X$ which can take possible values$x_m, m = 0, 1 \dots M$ where $x_0$ denotes the level at which the risk of the outcome is the lowest. The population attributable fraction for a binary outcome $Y$, where $Y = 1$ denotes the detrimental event, is given by

$$PF = \frac{\Pr(Y=1) - Pr_c\,(Y=1|X=x_0)}{\Pr\,(Y=1)}, \tag{1}$$

where $\Pr\,(Y = 1)$ denotes the probability of the outcome in a current population and $Pr_c\,(Y = 1|X = x_0)$ denotes the probability of the outcome in a counterfactual population where the exposure level of all individuals is shifted to $x_0$, *without changing the distribution of any other co-factors*.

In the absence of any other co-factors, the PF can be conveniently represented in the form[1-3] ,

$$PF = 1 - \; 1/\sum_{m=0}^{M}\{RR_m \times \Pr\,(X = x_m)\} \tag{2}$$

where $RR_m$ denote the associated with the exposure category $X = x_m$ relative to $X = x_0$ with the convention $RR_0 = 1$. This representation of PF, however, is generally not valid in the presence of potential confounders and effect modifiers. In the presence of known co-factors $(Z)$, one can develop a model for $\Pr\,(Y|X, Z)$ based on observed data and then derive the required counterfactual probability $Pr_c\,(Y = 1|X = x_0)$ as

$$Pr_c(Y|X = x_0) = \int \Pr(Y|X = x_0, Z)\, dF(Z) \tag{3}$$

where $F(Z)$ denotes the distribution of the co-factors in the underlying target population.

Using an underlying log-linear model for confounders (measure or unmeasured), we derive an expression for $Pr_c(Y = 1|X = x_0)$ as (see Section 2)

$$Pr_c(Y = 1|X = x_0) = \Pr(Y = 1|X = x_0) \times \left\{ \sum_{m=0}^{M} \frac{RR_m^U}{RR_m^A} \Pr(X = x_m) \right\} \quad (4)$$

where $\Pr(Y = 1|X = x_0)$ denotes the observed probability of the outcome in the baseline exposure category and $RR_m^U$ and $RR_m^A$ , $m = 1,..M$ denote the unadjusted/raw and adjusted estimates of relative risk parameters associated with the exposure categories. Using (4) and (1), we derive an alternative formula for PF as

$$PF = 1 - \frac{\Pr(Y = 1|X = x_0) \times \left\{ \sum_{m=0}^{M} \frac{RR_m^U}{RR_m^A} \Pr(X = x_m) \right\}}{\Pr(Y = 1)} \quad (5)$$

We further show in section 2 that the same formula can be derived based on an alternative representation of PF that has been proposed requiring estimates of exposure prevalence from representative cases.

According to representation (4), we can show the numerator of PF (1) can be decomposed as

$$\Pr(Y = 1) - Pr_c(Y = 1|X = x_0)$$
$$= \sum_{m=1}^{M} [\Pr(Y = 1|X = x_m) - \{\Pr(Y = 1|X = x_0) \times \tfrac{RR_m^U}{RR_m^A}\}] \Pr(X = x_m), \quad (6)$$

where each term in the sum is proportional to the number of attributable events from the corresponding category of the exposure. In the above, we can further show that $Pr(Y = 1|X = x_0) \times \frac{RR_m^A}{RR_m^U}$ corresponds to the counterfactual probability of the outcome in a sub-population whose exposure value has been intervened to change from $x_m$ to $x_0$. Thus, we define the quantity

$$Pr_c^{x_m \to x_0}(Y = 1|X = x_0) = Pr(Y = 1|X = x_0) \times \frac{RR_m^A}{RR_m^U}, \quad (7)$$

and note that the quantity $RD_c^{x_m \to x_0} = \{\Pr(Y = 1|\, X = x_m) - Pr_c^{\, x_m \to x_0}(Y = 1|X = x_0)\}$ denote individual-level average risk reduction due to modification of exposure from $x_m$ to $x_0$. We can now further define PF associated with shifting of BMI distribution in a population from the observed $(F_O)$ to any hypothetical distribution $(F_H)$ as follows. Let $p_{\{x_m \to x_{m'}\}}, m = 1,..M; m' = 1, \dots M;$ denote the transition probabilities associated with shifting of the BMI distribution from the observed to the hypothetical distribution (see e.g., Supplemental Tables 7 and 8 for transition probabilities corresponding to excess BMI reduction of 50% across the board in the NHANES population). We can define the probability of death in the resulting counterfactual population as

$$Pr_{F_H}(Y = 1) = \sum_{m=0}^{M} \sum_{m'=0}^{M} \Pr(Y = 1|X = x_{m'}) \frac{RR_m^U}{RR_m^A} \times \frac{RR_{m'}^A}{RR_{m'}^U}\, p_{\{x_m \to x_{m'}\}} \quad (8)$$

and the corresponding PF can be defined using formula (1) and is given by,

$$PF^{F_o \to F_H} = 1 - \frac{\sum_{m=0}^{M} \sum_{m'=0}^{M} \Pr(Y=1|X=x_{m'}) \frac{RR_m^U}{RR_m^A} \times \frac{RR_{m'}^A}{RR_{m'}^U} p_{\{x_m \to x_{m'}\}}}{\Pr(Y=1)} \quad (9)$$

Finally, we can show that the formulae (4), (6), (7), (8) and (9) can be easily generalized conditional on any observed co-factor value $S = s$ by replacing throughout $\Pr(X = x_m)$ by $\Pr(X = x_m | S = s)$, and $RR_m^A$ and $RR_m^U$ by corresponding adjusted/stratified effects. We observe that external results, such as those from MR studies, may not provide estimates of the effects of the exposure stratified by other co-factors and thus one will need to make an implicit assumption of homogeneity of causal effects (in relative-risk scale). One can, however, estimate the observable relative risks($RR_m^U$) from a study like NHANES stratified by other measured co-factors using either empirical or modeling approaches. In our analysis, we define strata ($S$) based on quintiles of a risk score for mortality defined by all other co-factors considered. We used a modeling approach to define strata where we adjusted for other co-factors in the model (details provided in section 4). We did not explore interactions due to a low number of events. The total PF can be partitioned into a sum of contributions across strata as

$$PF = \frac{\sum_s \{\Pr(Y = 1|S = s) - Pr_C(Y = 1|X = x_0, S = s)\} \Pr(S=s)}{\Pr(Y=1)} \quad (10)$$

and thus, each term of the numerator as a ratio of the total provides quantification of the proportional contribution of each stratum to the total preventable death.

### 2. Derivation of PF formula (4) and its equivalence with Bruzzi's formula

#### a. Derivation of PF formula (4)

We assume the outcome model to take the following log-linear form

$$P(Y = 1|X, W) = \; exp\big(\beta_0 + \textstyle\sum_{m=1}^{M-1} \beta_{E,m} \mathbb{I}_{\{X=x_m\}} + W\big) \quad (11)$$

where $W \coloneqq \sum_{k=1}^{K} \beta_k W_k$ is a weighted linear combination of all possible, measured and unmeasured, $K$ confounders denoted by $W_1, \dots W_K$. We further assume the conditional distribution of $W|X = x_m$ follows a normal distribution with mean $\mu_m$ and variance $\sigma_m^2$ for $m = 0, \dots, M-1$. Then, we have

$$Pr_C(Y = 1|X = x_0) \; = \int P(Y = 1|X = x_0, W = w) dF(w)$$

$$= \int P(Y = 1|X = x_0, W = w) \left\{ \sum_{m=0}^{M-1} dF_{W|X}(w|X = x_m) P(X = x_m) \right\}$$

$$= \frac{1}{\sqrt{2\pi}} \exp\left(\beta_0\right) \sum_{m=0}^{M-1} P(E = m) \frac{\exp\left(\mu_m + \sigma_m^2/2\right)}{\sigma_m} \quad (12)$$

Using the model (11), the observed probability of the outcome at the baseline category can be derived as

$$P(Y = 1|X = x_0) = \int P(Y = 1|X = x_0, W = w) dF_{W|X}(w|X = x_0)$$

$$= \frac{1}{\sqrt{2\pi}} \exp\left(\beta_0\right) \frac{\exp\left(\mu_0 + \sigma_0^2/2\right)}{\sigma_0} \quad (13)$$

Note that the unadjusted relative risk, denoted by $RR_m^U, m = 1, \dots, M-1$, can be derived from model (1) and is given by

$$RR_m^U = \frac{\Pr\left(Y=1|X=x_m\right)}{\Pr(Y=1\ |X=x_0)} = \exp\left(\beta_m\right) \frac{\exp\left(\mu_m + \sigma_m^2/2\right)/\sigma_m}{\exp\left(\mu_0 + \sigma_0^2/2\right)/\sigma_0} \quad (14)$$

Using (12), (13) and (14), we have the following formula (formula (4))

$$Pr_C(Y = 1|X = x_0) = Pr(Y = 1|X = x_0) \left\{ \sum_{m=0}^{M-1} \frac{RR_m^U}{RR_m^A} Pr(X = x_m) \right\}$$

where, $RR_m^A = \exp\left(\beta_m\right)$ is the adjusted relative risk of exposure category $m$, for $m = 1, \dots, M-1$, and $RR_0^U = RR_0^M = 1$. We evaluate, $\beta_m$, by directly plugging in the value $\widehat{\beta_m^{MR}}$ from an external MR-based study.

### b. Equivalence with Bruzzi's formula

Note that by definition, $RR_m^U = \frac{\Pr\left(Y=1|X=x_m\right)}{\Pr\left(Y=1|X=x_0\right)}$. Then, using our proposed formula (5), we have

$$PF = 1 - \frac{\Pr(Y = 1|X = x_0) \left\{ \Pr(X = x_0) + \sum_{m=1}^{M-1} \frac{RR_m^U}{RR_m^A} \Pr(X = x_m) \right\}}{\Pr(Y = 1)}$$

$$= \frac{\{\Pr(Y = 1) - \Pr(Y = 1, X = x_0)\} - \sum_{m=1}^{M-1} \Pr(Y = 1|X = x_0) \frac{\Pr(Y = 1|X = x_m)}{\Pr(Y = 1|X = x_0) RR_m^A} \Pr(X = x_m)}{\Pr(Y = 1)}$$

$$= \frac{\sum_{m=1}^{M-1} \Pr(Y=1, X=x_m) - \sum_{m=1}^{M-1} \Pr(Y=1, X=x_m) \frac{1}{RR_m^A}}{\Pr(Y=1)}$$

$$= \sum_{m=0}^{M-1} \Pr(X = x_m \mid Y = 1)\left\{1 - \frac{1}{RR_m^A}\right\}$$

$$= \; PF^{Bruzzi}$$

where $PF^{Bruzzi}$ is the formula provided by Bruzzi[3,4], and $RR_0^A = 1$.

#### 3. Variance formula

All the variance formulae are evaluated using the Delta method[5]. From now on, we will denote an estimate of any quantity, $A$ by $\hat{A}$. Let us denote $\widehat{\Pr(X = x_m)}$ by $\widehat{p_m}$ and $\widehat{\Pr(Y = 1)}$ by $\hat{p}$. We estimate the quantity, $\widehat{RR_m^U}$, by $\exp\widehat{(\theta_m)}$, for $m = 1, \dots M-1$, and $\widehat{\Pr(Y = 1|X = x_0)}$ by $\exp\widehat{(\theta_0)}$, where $\widehat{\theta_m}$ is the maximum likelihood estimate based on the following logistic regression model,

$$P(Y = 1|X) = expit(\theta_0 + \textstyle\sum_{m=1}^{M} \theta_m \mathbb{I}_{\{X=x_m\}}) \tag{15}$$

. Note that all the estimates so far are being estimated from the study sample underlying the target population. Any formula of PF described in section 1 can be written as some non-linear vector to scalar function, $g(\widehat{\gamma_1}, \widehat{\gamma_2})$, where $\widehat{\gamma_1} = \left(\widehat{\theta_0}, \widehat{\theta_1}, \dots, \widehat{\theta_{M-1}}, \widehat{p_1}, \dots, \widehat{p_{M-1}}, , \hat{p}\right)$ and $\widehat{\gamma_2} = \left(\widehat{\log RR_1^A}, \dots, \widehat{\log RR_{M-1}^A}\right)$. Using delta method, we have the following general formula for calculating the variance of $\widehat{PF}$

$$Var\widehat{(PF)} = \; \widehat{\boldsymbol{a}}^T \widehat{\boldsymbol{\Sigma}} \widehat{\boldsymbol{a}} + \widehat{\boldsymbol{b}}^T \; \widehat{\boldsymbol{\Omega}} \widehat{\boldsymbol{b}} \tag{16}$$

where, $\widehat{\boldsymbol{\Sigma}}$ is the variance-covariance matrix of $\widehat{\gamma_1}^T$, $\widehat{\boldsymbol{\Omega}}$ is the variance-covariance matrix of the adjusted relative risk vector, $\widehat{\gamma_2}^T$ from the external MR-based study, $\widehat{\boldsymbol{a}} = \left(\frac{\partial g}{\partial \gamma_1} \mid_{\gamma_1 = \widehat{\gamma_1}}\right)^T$ and $\widehat{\boldsymbol{b}} = \left(\frac{\partial g}{\partial \gamma_2} \mid_{\gamma_2 = \widehat{\gamma_2}}\right)^T$ . Intuitively, formula (16) can be seen as a sum of two independent terms. The first term accounts for the uncertainty arising from the internal study sample based on which the exposure prevalences, the proportion of deaths, baseline absolute risk, and unadjusted relative risk are estimated. The second term accounts for the uncertainty arising from an external randomized study.

### 4. Data Analysis Steps

We downloaded the NHANES 1999-2006 and 2017-18 datasets from the Centers for Disease Control and Prevention (CDC) online portal[6]. We also downloaded the linked mortality data till 2015 from National Death Index for each of the biennial cycles of the NHANES 1999-2006 cohort. We obtained the MR-based estimates of relative risks of WHO-defined BMI categories ($x_m/x_{m'}$) on mortality from Sun et al. From table 1, we have 22.06 – 25.66 Kgm$^{-2}$ ($x_0$), 25.66 – 27.78 Kgm$^{-2}$ ($x_1$), 27.78 – 30.73 Kgm$^{-2}$ ($x_2$), 30.73 – 36.13 Kgm$^{-2}$ ($x_3$), 36.13 – 42.35 Kgm$^{-2}$ ($x_4$), ≥ 42.35 Kgm$^{-2}$ ($x_5$) as the six BMI categories for the entire cohort.

We restricted the analysis to normal and overweight individuals with age 40-69 years. The cause-specific mortality outcomes ($Y$) are defined according to the underlying leading cause of death which is coded as 001 for diseases of the heart and 002 for malignant neoplasms. These codes are according to the manual provided with the mortality data. We analyzed the data separately for the non-Hispanic White and non-Hispanic Black populations.

We estimated the prevalence of each BMI category empirically by taking a weighted mean of the individuals in each of the BMI categories, where weights are the sampling weights provided by NHANES. The estimates are shown in Table 1 for the non-Hispanic White population and Supplementary Table 3 for the non-Hispanic Black population. Similarly, we estimated the prevalence of all the mortality outcomes for NHANES 1999-2006. We used model-based approaches to evaluate other components in the PF formulas. All the models that we considered in the analysis are logistic regression models. The unadjusted/raw relative risk estimates of BMI categories ($RR_m^U$) are estimated using the odd-ratio estimates of the BMI categories obtained from the model (15) with BMI as the categorical regressor and mortality as the outcome. The adjusted relative risk estimates of BMI categories ($RR_m^A$) are estimated using the MR-based estimates of relative risks. For the non-Hispanic Black population, we used the same estimates for adjusted relative risk assuming the MR estimates of relative risks to be the same as that of the White population. The estimates for the unadjusted/raw and adjusted relative risk estimates for the non-Hispanic White and Black populations are shown in Table 1 and Supplementary Table 3, respectively. We plugged in these estimates for the non-Hispanic White/non-Hispanic Black populations along with the prevalence of BMI categories for NHANES 1999-2006 from Table 1/Supplementary Table 3 and prevalence of the mortality outcome of interest in formula (5) to obtain the estimates of $PF_{100\%}$ shown in Table 1/Supplementary Table 4. We plugged in the same empirical and model-based estimates in formula (9) for evaluating $PF_{50\%}$. The transition probabilities in formula (9) are estimated empirically and are shown in supplementary table 7/8 for the non-Hispanic White/non-Hispanic Black population. The results for $PF_{50\%}$ are shown in Table 1/Supplementary Table 4 for the non-Hispanic White/non-Hispanic Black population. In the results shown in supplementary table 5, where we calculated $PF_{100\%}$ and $PF_{50\%}$ for the non-Hispanic White population using formulas (5) and (9) respectively, we plugged in the relative risk estimates based on external pooled study[7] (shown in Table 1) for the adjusted relative risk estimate of BMI categories ($RR_m^A$), instead of the MR-based estimates. All the other components are evaluated using the same estimates as before from Table 1.

We calculated the projected PF for the NHANES 2017-18 assuming the unadjusted relative risk ($RR_m^U$) of BMI categories on the risk of mortality to be the same across NHANES 1999-2006 and 2017-18. Under this assumption, we estimated the prevalence of the outcome for NHANES 2017-18, $\Pr(Y=1)$ by $\sum_{m=0}^{M} \Pr(Y=1|X=x_m)\Pr(X=x_m)$, where $\Pr(Y=1|X=x_m)$ is estimated based on a logistic regression model (14) fit to the NHANES 1999-2006 data, and $\Pr(X=x_m)$ is estimated empirically from the NHANES 2017-18 data. Using $\Pr(Y=1) = \sum_{m=0}^{M} \Pr(Y=1|X=x_m)\Pr(X=x_m)$ in the denominator of PF formulae (5) and (9), we obtain the following corresponding projected PF formulae,

$$PF_{projected} = 1 - \frac{\sum_{m=0}^{M} \frac{RR_m^U}{RR_m^A} \Pr(X=x_m)}{\sum_{m=0}^{M} RR_m^U \Pr(X=x_m)} \quad (17)$$

$$PF_{projected}^{F_O \to F_H} = 1 - \frac{\sum_{m=0}^{M} \sum_{m'=0}^{M} \Pr(Y=1|X=x_{m'}) \frac{RR_m^U}{RR_m^A} \times \frac{RR_{m'}^A}{RR_{m'}^U} p_{\{x_m \to x_{m'}\}}}{\Pr(Y=1|X=x_0)\{\sum_{m=0}^{M} RR_m^U \Pr(X=x_m)\}} \quad (18)$$

where $p_{\{x_m \to x_{m'}\}}$ are the transition probabilities as mentioned earlier in section 1, formula (9). We evaluated the projected $PF_{100\%}$ and $PF_{50\%}$ for NHANES 2017-18 using formulae (17) and (18) respectively. The results are shown in Table 2 and Supplementary Table 4. The components, $RR_m^U$ and $RR_m^A$ in formulae (17), and (18) are evaluated using the same estimates as mentioned earlier for NHANES 1999-2006. Conditional probabilities, $\Pr(Y=1|X=x_{m'})$, and $\Pr(Y=1|X=x_0)$ are estimated based on model (14) fit to the NHANES 1999-2006. The transition probabilities (supplementary tables 7/8 for the non-Hispanic White/non-Hispanic Black population) and the prevalence of BMI categories for NHANES 2017-18 (table 1/supplementary table 3 for the non-Hispanic White/non-Hispanic Black population) are estimated empirically from the NHANES 2017-18, instead of NHANES 199-2006, using weighted sample means where weights are the sample weights provided in NHANES 2017-18.

We defined the risk score of an individual as a weighted combination of the individual's risk factors other than BMI. The weights are the estimated odds-ratio (in log-scale) parameters of the logistic regression model fit to the NHANES 1999-2006 data. The risk factors included in the model are age, sex, education, smoking status, average number of cigarettes smoked per day, alcohol status, and marital status. We computed the absolute risk reduction associated with a reduction in BMI by 100% and 50% (Figure 1 and supplementary figures 1-5) for quintiles of risk score using extending the formula (8) as mentioned in section 1, by conditioning on quintile. Here, $RR_m^U$ is evaluated as the odds ratio estimate of the BMI category m, adjusted for all other risk factors (used to define the risk score) under a logistic regression model. The quantity, $\Pr(Y=1|X=x_{m'}, quintile\ of\ risk\ score)$, is evaluated based on a logistic regression model. We first developed the model using all the risk factors including BMI. Using the model, we then evaluated the risk of dying for an individual who belongs to the BMI category, $x_{m'}$, and a particular quintile of risk score. Finally, we took the average of the risk across individuals belonging to that BMI and quintile category. The rest of the quantities, quintile-specific BMI category prevalence, and prevalence of disease are evaluated empirically from the data.

We also partitioned the total PF ($PF_{100\%}$) due to excess BMI over broader overweight categories, pre-obese and severe obese, across quintiles of risk scores defined earlier. The results are shown in Table 3. The estimates are calculated based on the data on NHANES 1999-2006 non-Hispanic White population using formula (10) where strata here is defined according to combined categories of the overweight categories and the quintiles of risk score defined before. For each of the strata, we first calculated the numerator of formula (10) as a ratio of the total ($\Pr(Y = 1)$) for each term corresponding to each category, and finally estimated the contribution from each category as a ratio of the total PF($PF_{100\%}$). The quantities needed for formula (10), $RR_m^U$ and $\Pr(Y = 1|X = x_m, quintile\ of\ risk\ score)$ are estimated in the same way as described in the earlier paragraph, but with the broader BMI categories in the logistic regression model. For estimating the adjusted relative risk estimate in formula (10), $RR_m^A$ , we assumed a linear relationship of BMI on mortality for the broader categories and evaluated the average effect within each of the broader overweight categories from the external MR-based study. The resulting estimates are plugged-in for $RR_m^A$ .
