## Supplementary Tables and Figures for "Estimating Burden of Mortality due to Excess Body Mass Index in the US Adult Population by Combining Evidence from a Mendelian Randomization Study and National Health Surveys"

***Supplementary Table 1: Characteristics of the National Health and Nutritional Examination Survey (NHANES) <u>non-Hispanic White participants</u> from 1999 to 2006.*** *The values shown in this table are based on unweighted analysis.*

| | BMI percentile-based category in $Kgm^{-2}$ | | | | | |
|---|---|---|---|---|---|---|
| | **22.1 – 25.7** | **25.7 – 27.7** | **27.7 – 30.7** | **30.7 – 36.1** | **36.1 – 42.3** | **≥ 42.3** |
| **Total sample size** | 853 | 596 | 745 | 812 | 346 | 139 |
| **Number (%) of deaths from all cause** | 101 (11.8) | 82 (13.8) | 102 (13.7) | 117 (14.4) | 66 (19.1) | 26 (18.7) |
| **from heart-diseases** | 61 (2.6) | 49 (3.4) | 60 (3.7) | 52 (3.3) | 24 (3.9) | 9 (3.6) |
| **from cancers** | 91 (3.8) | 69 (4.8) | 82 (5.0) | 68 (4.3) | 31 (5.0) | 8 (3.2) |
| **Mean age (SD)** | 53.15 (8.68) | 54.64 (8.79) | 54.50 (8.75) | 54.41 (8.58) | 54.92 (8.29) | 53.02 (8.65) |
| **Number (%) of males** | 1164 (49.0) | 774 (54.3) | 957 (58.6) | 798 (50.7) | 235 (37.8) | 87 (34.9) |
| **Number (%) by smoking status** | | | | | | |
| **Current** | 235 (27.5) | 130 (21.8) | 152 (20.4) | 162 (20.0) | 67 (19.4) | 22 (15.8) |
| **Former** | 246 (28.8) | 213 (35.7) | 285 (38.3) | 298 (36.7) | 135 (39.0) | 47 (33.8) |
| **Never** | 372 (43.6) | 253 (42.4) | 308 (41.3) | 352 (43.3) | 144 (41.6) | 70 (50.4) |
| **Average no. of cigarettes smoked per day (SD)** | 11.25 (14.30) | 10.90 (14.15) | 12.82 (16.26) | 12.68 (16.50) | 13.75 (17.34) | 11.85 (17.96) |
| **Number (%) by alcohol status** | | | | | | |
| **Heavy** | 90 (11.0) | 65 (11.4) | 74 (10.4) | 58 (7.5) | 15 (4.6) | 1 (0.7) |
| **Moderate** | 532 (64.9) | 356 (62.3) | 446 (62.6) | 449 (57.8) | 166 (51.1) | 72 (52.9) |
| **Non-drinker** | 198 (24.1) | 150 (26.3) | 193 (27.1) | 270 (34.7) | 144 (44.3) | 63 (46.3) |
| **Number (%) by education status** | | | | | | |
| **College graduate or above** | 293 (34.4) | 193 (32.4) | 212 (28.5) | 190 (23.4) | 74 (21.4) | 25 (18.0) |
| **Some college or AA degree** | 238 (27.9) | 164 (27.5) | 232 (31.2) | 249 (30.7) | 115 (33.2) | 46 (33.1) |
| **High School Grad/GED or equivalent** | 223 (26.2) | 163 (27.3) | 205 (27.6) | 240 (29.6) | 97 (28.0) | 49 (35.3) |
| **9-11th Grade (Includes 12th grade with no diploma)** | 75 (8.8) | 52 (8.7) | 67 (9.0) | 98 (12.1) | 44 (12.7) | 15 (10.8) |
| **Less than 9th Grade** | 23 (2.7) | 24 (4.0) | 28 (3.8) | 34 (4.2) | 16 (4.6) | 4 (2.9) |
| **Number (%) by marital status** | | | | | | |
| **Divorce** | 177 (22.7) | 111 (19.7) | 136 (19.5) | 149 (19.3) | 74 (22.6) | 30 (22.6) |
| **Married** | 566 (72.7) | 428 (76.0) | 523 (75.0) | 588 (76.1) | 232 (70.9) | 81 (60.9) |
| **Never married** | 36 (4.6) | 24 (4.3) | 38 (5.5) | 36 (4.7) | 21 (6.4) | 22 (16.5) |

***Supplementary Table 2: Characteristics of the National Health and Nutritional Examination Survey (NHANES) <u>non-Hispanic Black</u> participants from 1999 to 2006 for all-cause mortality.*** *The values shown in this table are based on unweighted analysis.*

| | BMI percentile-based category in $Kgm^{-2}$ | | | | | |
|---|---|---|---|---|---|---|
| **Variables** | **22.1 – 25.7** | **25.7 – 27.7** | **27.7 – 30.7** | **30.7 – 36.1** | **36.1 – 42.3** | **≥ 42.3** |
| **Total sample size** | 329 | 266 | 343 | 395 | 222 | 133 |
| **Number (%) of deaths from all cause** | 66 (20.1) | 41 (15.4) | 51 (14.9) | 68 (17.2) | 37 (16.7) | 23 (17.3) |
| **Age (mean (SD))** | 53.16 (8.72) | 52.72 (8.59) | 53.74 (8.97) | 53.49 (9.08) | 54.37 (9.00) | 53.02 (8.83) |
| **Number (%) of males** | 211 (64.1) | 147 (55.3) | 164 (47.8) | 174 (44.1) | 74 (33.3) | 30 (22.6) |
| **Number (%) by smoking status** | | | | | | |
| **Current** | 128 (39.0) | 80 (30.2) | 88 (25.7) | 82 (20.8) | 43 (19.5) | 22 (16.5) |
| **Former** | 72 (22.0) | 59 (22.3) | 78 (22.7) | 111 (28.1) | 60 (27.1) | 29 (21.8) |
| **Never** | 128 (39.0) | 126 (47.5) | 177 (51.6) | 202 (51.1) | 118 (53.4) | 82 (61.7) |
| **Average no. of cigarettes smoked per day (mean (SD))** | 8.75 (11.06) | 7.23 (10.44) | 6.15 (10.17) | 7.68 (12.77) | 6.78 (10.78) | 5.80 (10.90) |
| **Number (%) by alcohol status** | | | | | | |
| **Heavy** | 26 (8.8) | 19 (7.9) | 22 (7.0) | 15 (4.1) | 6 (2.9) | 2 (1.6) |
| **Moderate** | 156 (52.5) | 131 (54.1) | 156 (49.4) | 184 (50.5) | 79 (38.5) | 53 (41.4) |
| **Non-drinker** | 115 (38.7) | 92 (38.0) | 138 (43.7) | 165 (45.3) | 120 (58.5) | 73 (57.0) |
| **Number (%) by education status** | | | | | | |
| **College graduate or above** | 52 (15.9) | 48 (18.1) | 62 (18.1) | 62 (15.7) | 23 (10.4) | 19 (14.3) |
| **Some college or AA degree** | 83 (25.3) | 71 (26.8) | 95 (27.8) | 119 (30.1) | 70 (31.5) | 42 (31.6) |
| **High School Grad/GED or equivalent** | 75 (22.9) | 63 (23.8) | 72 (21.1) | 105 (26.6) | 47 (21.2) | 27 (20.3) |
| **9-11th Grade (Includes 12th grade with no diploma)** | 88 (26.8) | 67 (25.3) | 94 (27.5) | 87 (22.0) | 66 (29.7) | 36 (27.1) |
| **Less than 9th Grade** | 30 (9.1) | 16 (6.0) | 19 (5.6) | 22 (5.6) | 16 (7.2) | 9 (6.8) |
| **Number (%) by marital status** | | | | | | |
| **Divorce** | 94 (32.9) | 85 (35.0) | 113 (36.0) | 123 (33.5) | 88 (42.5) | 52 (43.0) |
| **Married** | 137 (47.9) | 121 (49.8) | 163 (51.9) | 185 (50.4) | 90 (43.5) | 44 (36.4) |
| **Never married** | 55 (19.2) | 37 (15.2) | 38 (12.1) | 59 (16.1) | 29 (14.0) | 25 (20.7) |

***Supplementary Table 3: Population prevalence and risk of mortality (95% CI) associated with BMI categories for the <u>non-Hispanic Black population</u>.*** *Prevalence of BMI categories are derived from the NHANES 1999-2006/2017-18 data. Relative-risks of mortality for BMI categories are calculated with respect to the reference category 22.06 – 25.66 Kgm$^{-2}$. Two sets of relative-risk parameters correspond to (1) internal analysis of NHANES 1999-2006 data linked to all-cause mortality; and (2) external Mendelian randomization study based on the White population.*

| **BMI categories** | **25.66 – 27.78** | **27.78 – 30.76** | **30.76 – 36.13** | **36.13 – 42.35** | **≥ 42.35** |
|---|---|---|---|---|---|
| **Prevalence of the BMI categories in NHANES[1] 1999-2006 /2017-18(in %)** | 16/14 | 20/17 | 23/27 | 13/17 | 8/12 |
| | | | **Odds ratio for all-cause mortality** | | |
| **NHANES[1] (95% CI)** | 0.76 (0.48 – 1.21) | 0.76 (0.46 – 1.26) | 0.84 (0.51 – 1.37) | 0.78 (0.47 – 1.30) | 0.94 (0.56 – 1.58) |
| **MR estimate[2] (95% CI)** | 1.05 (0.99 – 1.11) | 1.20 (1.05 – 1.36) | 1.74 (1.26 – 2.41) | 3.49 (1.83 – 6.66) | 5.95 (2.57 – 13.78) |

[1] *All analyses of NHANES data incorporated sample weights. The odds ratios from the NHANES analysis are not adjusted for any other factors and these "raw" estimates are incorporated into PF calculations according to formula (5) of Supplemental Methods. See Section 4 of Supplemental Methods for details.*

[2] *The MR-estimates for the non-Hispanic Black population are assumed to be same as that of the White population from the external MR-based study[25].*

***Supplementary Table 4: Estimates (in %) of Preventable Fraction (PF) for 10-year mortality in self-reported <u>non-Hispanic Black</u> individuals and age group 40-69 in the NHANES populations for the time periods 1999-2006 and 2017-18****. Two sets of estimates of PF are obtained corresponding to 100% ($PF_{100\%}$)[1] and 50% ($PF_{50\%}$)[2] reduction of excess BMI (compared to normal weight) across all individuals in the underlying populations. Results are derived using BMI prevalence data from NHANES, linked mortality outcome data for the 1999-2006 NHANES cohort, and estimates of BMI effects from an external MR-study. The reference categories for BMI are chosen according to those provided by the MR-study: 22.06 – 25.66 $Kgm^{-2}$ for the whole population, 22.3 – 26 $Kgm^{-2}$ for ever smokers and 21.9 – 25.4 $Kgm^{-2}$ for never smokers. All analyses of NHANES data incorporate sampling weights.*

| Mortality outcome | Smoking status | Estimate (in %) of $PF_{100\%}$ (95% CI) | | Estimate (in %) of $PF_{50\%}$ (95% CI) | |
|---|---|---|---|---|---|
| | | 1999-2006 | Projected for 2017-18 | 1999-2006 | Projected for 2017-18 |
| All-cause | All | 35 (19 – 50) | 37 (24 – 51) | 22 (11 – 32) | 26 (16 – 36) |
| | Ever [a] | 30 (14 – 46) | 35 (19 – 51) | 19 (4 – 33) | 25 (14 – 36) |
| | Never | 32 (-4 – 68) | 36 (-1 – 74) | 19 (-9 – 47) | 20 (-8 – 48) |

[1] *$PF_{100\%}$ for 1999-2006 and for 2017-18 are respectively calculated using the formula (5) and formula (17) provided in the supplementary methods. See Section 4 of Supplemental Methods for details.*

[2] *$PF_{50\%}$ for 1999-2006 using formula (9) of supplementary methods. $PF_{50\%}$ projected for 2017-18 is calculated using formula (18) provided in the supplementary methods. See Section 4 of Supplemental Methods for details.*

[a] *Ever smokers include former and current smokers.*

***Supplementary Table 5: Estimates (in %) of Preventable Fraction (PF) for 10-year mortality in self-reported non-Hispanic White individuals and age group 40-69 in the NHANES population** for the time period 1999-2006. Two sets of estimates of PF are obtained corresponding to 100% ($PF_{100\%}$)*[a] *and 50% ($PF_{50\%}$)*[b] *reduction of excess BMI (compared to normal weight) across all individuals in the underlying populations. Results are derived using BMI prevalence data from NHANES, linked mortality outcome data for the 1999-2006 NHANES cohort, and <u>estimates of BMI effects from external pooled-cohort analysis</u>*[c]*. The reference category for BMI is chosen according to one provided by the MR-study: 22.06 – 25.66 $Kgm^{-2}$ for the whole population. All analyses of NHANES data incorporate sampling weights.*

| Race | Mortality outcome | Estimate (in %) of $PF_{100\%}$ (95% CI[d]) | | Estimate (in %) of $PF_{50\%}$ (95% CI[d]) | |
|---|---|---|---|---|---|
| | | **1999-2006** | **Projected for 2017-18** | **1999-2006** | **Projected for 2017-18** |
| **White** | All-cause | 22 (16 – 29) | 26 (20 – 32) | 12 (-1 – 26) | 18 (14 – 22) |
| | Heart disease | 36 (28 – 45) | 41 (33 – 49) | 22 (-4 – 48) | 30 (23 – 37) |
| | Cancer | 16 (7 – 25) | 16 (9 – 23) | 8 (-11 – 28) | 12 (-9 – 33) |

[a] *$PF_{100\%}$ for 1999-2006 and for 2017-18 are respectively calculated using the formula (5) and formula (17) provided in the supplementary methods. See Section 4 of Supplemental Methods for details.*

[b] *$PF_{50\%}$ for 1999-2006 using formula (9) of supplementary methods. $PF_{50\%}$ projected for 2017-18 is calculated using formula (18).*

[c] *The pooled estimates are based on analysis of all Western cohorts as reported in Di Angelantonio, E., et.al [9]. The BMI categories used in this study are similar to the one used for the external MR study[25], but they are not identical. Note that the pooled estimates are based on analysis in never-smokers without known chronic disease at baseline and excluding the first 5 years of follow-up.*

[d] *It is assumed the off-diagonal elements of the variance-covariance matrix of the BMI effects from the external pooled-cohort analysis are zero.*

***Supplementary Table 6: Use of the widely used Walter's formula[a] to estimate PF (in %) for 10-year mortality due to excess BMI for the NHANES 1999-2006 non-Hispanic White population with age range 40-69****. The reference category for BMI is chosen according to one provided by the MR-study: 22.06 – 25.66 Kgm$^{-2}$ for the whole population. All analyses of NHANES data incorporate sampling weights.*

| Race | Mortality outcome | Using MR relative risk estimates (95% CI)[b] | Using pooled relative risk estimates (95% CI)[c] |
|---|---|---|---|
| **White** | All-cause | 40 (25 – 55) | 23 (16 – 30) |
| | Heart disease | 48 (32 – 64) | 35 (28 – 42) |
| | Cancer | 19 (-4 – 42) | 16 (7 – 25) |

[a] *The Walter's formula is formula (3) of Table 1 as reported in Rockhill B, et.al.[18]*

[b] *The PF estimates are using Walter's formula with relative-risk estimates from the external MR-bases study reported in Sun et al[25].*

[c] *The PF estimates are using Walter's formula with relative-risk estimates as the pooled estimates based on analysis of all Western cohorts as reported in Di Angelantonio, E., et.al [9].*

***Supplementary Table 7: Proportion (transition probabilities[1]) of non-Hispanic white participants moving from one BMI category to another due to reduction in BMI by 50%.*** *The reference categories for BMI are chosen according to those provided by the MR-study: 22.06 – 25.66 Kgm$^{-2}$ for the whole population, 22.3 – 26 Kgm$^{-2}$ for ever smokers and 21.9 – 25.4 Kgm$^{-2}$ for never smokers.*

| Original BMI categories ↓ | Shifted BMI categories ⟶ | | | | | |
|---|---|---|---|---|---|---|
| | **1999-2006 (for the whole population)** | | | | | |
| | **Normal** | **Overweight** | **Severe Obese-I** | **Severe Obese-II** | **Severe Obese-III** | **Total** |
| **Normal** | 0.244 | 0 | 0 | 0 | 0 | 0.244 |
| **Overweight** | 0.384 | 0 | 0 | 0 | 0 | 0.384 |
| **Severe Obese-I** | 0.079 | 0.154 | 0 | 0 | 0 | 0.233 |
| **Severe Obese-II** | 0 | 0.049 | 0.05 | 0 | 0 | 0.099 |
| **Severe Obese-III** | 0 | 0 | 0.016 | 0.018 | 0.006 | 0.04 |
| **Total** | 0.707 | 0.203 | 0.066 | 0.018 | 0.006 | 1 |
| | **1999-2006 (for ever smokers/never smokers)** | | | | | |
| | **Normal** | **Overweight** | **Severe Obese-I** | **Severe Obese-II** | **Severe Obese-III** | **Total** |
| **Normal** | 0.256/0.225 | 0/0 | 0/0 | 0/0 | 0/0 | 0.256/0.225 |
| **Overweight** | 0.393/0.377 | 0/0 | 0/0 | 0/0 | 0/0 | 0.393/0.377 |
| **Severe Obese-I** | 0.084/0.073 | 0.135/0.177 | 0/0 | 0/0 | 0/0 | 0.219/0.250 |
| **Severe Obese-II** | 0/0 | 0.057/0.043 | 0.042/0.056 | 0/0 | 0/0 | 0.099/0.099 |
| **Severe Obese-III** | 0/0 | 0/0 | 0.014/0.021 | 0.014/0.021 | 0.005/0.007 | 0.033/0.049 |
| **Total** | 0.733/0.675 | 0.192/0.220 | 0.056/0.077 | 0.014/0.021 | 0.005/0.007 | 1/1 |
| | **2017-18 (for the whole population)** | | | | | |
| | **Normal** | **Overweight** | **Severe Obese-I** | **Severe Obese-II** | **Severe Obese-III** | **Total** |
| **Normal** | 0.139 | 0 | 0 | 0 | 0 | 0.139 |
| **Overweight** | 0.116 | 0.187 | 0 | 0 | 0 | 0.303 |
| **Severe Obese-I** | 0 | 0.277 | 0 | 0 | 0 | 0.277 |
| **Severe Obese-II** | 0 | 0.040 | 0.129 | 0 | 0 | 0.169 |
| **Severe Obese-III** | 0 | 0 | 0.064 | 0.040 | 0.008 | 0.112 |
| **Total** | 0.255 | 0.504 | 0.193 | 0.040 | 0.008 | 1 |
| | **2017-2018 (for ever smokers/never smokers)** | | | | | |
| | **Normal** | **Overweight** | **Severe Obese-I** | **Severe Obese-II** | **Severe Obese-III** | **Total** |
| **Normal** | 0.213/0.199 | 0/0 | 0/0 | 0/0 | 0/0 | 0.213/0.199 |
| **Overweight** | 0.340/0.312 | 0/0 | 0/0 | 0/0 | 0/0 | 0.340/0.312 |
| **Severe Obese-I** | 0.066/0.053 | 0.165/0.193 | 0/0 | 0/0 | 0/0 | 0.231/0.246 |
| **Severe Obese-II** | 0/0 | 0.056/0.069 | 0.069/0.090 | 0/0 | 0/0 | 0.125/0.159 |
| **Severe Obese-III** | 0/0 | 0/0 | 0.035/0.040 | 0.035/0.034 | 0.021/0.010 | 0.091/0.084 |
| **Total** | 0.619/0.564 | 0.221/0.262 | 0.104/0.130 | 0.035/0.034 | 0.021/0.010 | 1/1 |

*[1]These transition probabilities are used in formulae (8), (9) and (18) of the Supplementary Methods for calculating $PF_{50\%}$ for 1999-2006 and projected $PF_{50\%}$ for 2017-18 in the non-Hispanic White population. See Section 4 of Supplemental Methods for details.*

***Supplementary Table 8: Proportion (transition probabilities[1]) of non-Hispanic Black participants moving from one BMI category to another due to reduction in BMI by 50%.*** *The reference categories for BMI are chosen according to those provided by the MR-study: 22.06 – 25.66 Kgm$^{-2}$ for the whole population, 22.3 – 26 Kgm$^{-2}$ for ever smokers and 21.9 – 25.4 Kgm$^{-2}$ for never smokers.*

| Original BMI categories ↓ | Shifted BMI categories ⟶ | | | | | |
|---|---|---|---|---|---|---|
| | **1999-2006 (for the whole population)** | | | | | |
| | **Normal** | **Overweight** | **Severe Obese-I** | **Severe Obese-II** | **Severe Obese-III** | **Total** |
| **Normal** | 0.195 | 0 | 0 | 0 | 0 | 0.195 |
| **Overweight** | 0.137 | 0.224 | 0 | 0 | 0 | 0.361 |
| **Severe Obese-I** | 0 | 0.234 | 0 | 0 | 0 | 0.234 |
| **Severe Obese-II** | 0 | 0.042 | 0.089 | 0 | 0 | 0.131 |
| **Severe Obese-III** | 0 | 0 | 0.052 | 0.024 | 0.003 | 0.079 |
| **Total** | 0.332 | 0.500 | 0.141 | 0.024 | 0.003 | 1 |
| | **1999-2006 (for ever smokers/never smokers)** | | | | | |
| | **Normal** | **Overweight** | **Severe Obese-I** | **Severe Obese-II** | **Severe Obese-III** | **Total** |
| **Normal** | 0.246/0.142 | 0/0 | 0/0 | 0/0 | 0/0 | 0.246/0.142 |
| **Overweight** | 0.148/0.114 | 0.210/0.249 | 0/0 | 0/0 | 0/0 | 0.358/0.363 |
| **Severe Obese-I** | 0/0 | 0.225/0.247 | 0/0 | 0/0 | 0/0 | 0.225/0.247 |
| **Severe Obese-II** | 0/0 | 0.032/0.048 | 0.082/0.100 | 0/0 | 0/0 | 0.114/0.148 |
| **Severe Obese-III** | 0/0 | 0/0 | 0.036/0.066 | 0.019/0.031 | 0.002/0.003 | 0.057/0.100 |
| **Total** | 0.394/0.256 | 0.467/0.544 | 0.118/0.166 | 0.019/0.031 | 0.002/0.003 | 1/1 |
| | **2017-18 (for the whole population)** | | | | | |
| | **Normal** | **Overweight** | **Severe Obese-I** | **Severe Obese-II** | **Severe Obese-III** | **Total** |
| **Normal** | 0.201 | 0 | 0 | 0 | 0 | 0.201 |
| **Overweight** | 0.331 | 0 | 0 | 0 | 0 | 0.331 |
| **Severe Obese-I** | 0.066 | 0.183 | 0 | 0 | 0 | 0.249 |
| **Severe Obese-II** | 0 | 0.051 | 0.079 | 0 | 0 | 0.130 |
| **Severe Obese-III** | 0 | 0 | 0.043 | 0.03 | 0.016 | 0.089 |
| **Total** | 0.598 | 0.234 | 0.122 | 0.03 | 0.016 | 1 |
| | **2017-2018 (for ever smokers/never smokers)** | | | | | |
| | **Normal** | **Overweight** | **Severe Obese-I** | **Severe Obese-II** | **Severe Obese-III** | **Total** |
| **Normal** | 0.179/0.098 | 0/0 | 0/0 | 0/0 | 0/0 | 0.179/0.098 |
| **Overweight** | 0.088/0.120 | 0.175/0.201 | 0/0 | 0/0 | 0/0 | 0.263/0.321 |
| **Severe Obese-I** | 0/0 | 0.298/0.272 | 0/0 | 0/0 | 0/0 | 0.298/0.272 |
| **Severe Obese-II** | 0/0 | 0.042/0.049 | 0.126/0.128 | 0/0 | 0/0 | 0.168/0.177 |
| **Severe Obese-III** | 0/0 | 0/0 | 0.060/0.068 | 0.025/0.053 | 0.007/0.011 | 0.092/0.132 |
| **Total** | 0.267/0.218 | 0.515/0.522 | 0.186/0.196 | 0.025/0.053 | 0.007/0.011 | 1/1 |

*[1]These transition probabilities are used in formulae (8), (9) and (18) of the Supplementary Methods for calculating $PF_{50\%}$ for 1999-2006 and projected $PF_{50\%}$ for 2017-18 in the non-Hispanic Black population. See Section 4 of Supplemental Methods for details.*

**Supplementary Figure 1: Observed and counterfactual absolute risks of 10-year mortality due to heart-disease for obese individuals in the non-Hispanic White population with an age range of 40-69 represented by the NHANES 1999-2006 Surveys**. Observed risks correspond to empirical proportions of deaths observed. Counterfactual risks correspond to average of the estimated absolute risks of the individuals associated with 50% and 100% reduction of excess BMI (compared to normal weight) where MR-derived estimates of relative-risks are used to represent underlying counterfactual effects. Results are shown for the overall population and stratified by the quintile of a risk score defined by other prominent risk factors of mortality. See Sections 1 and 4 of Supplemental Methods for details of the derivations.

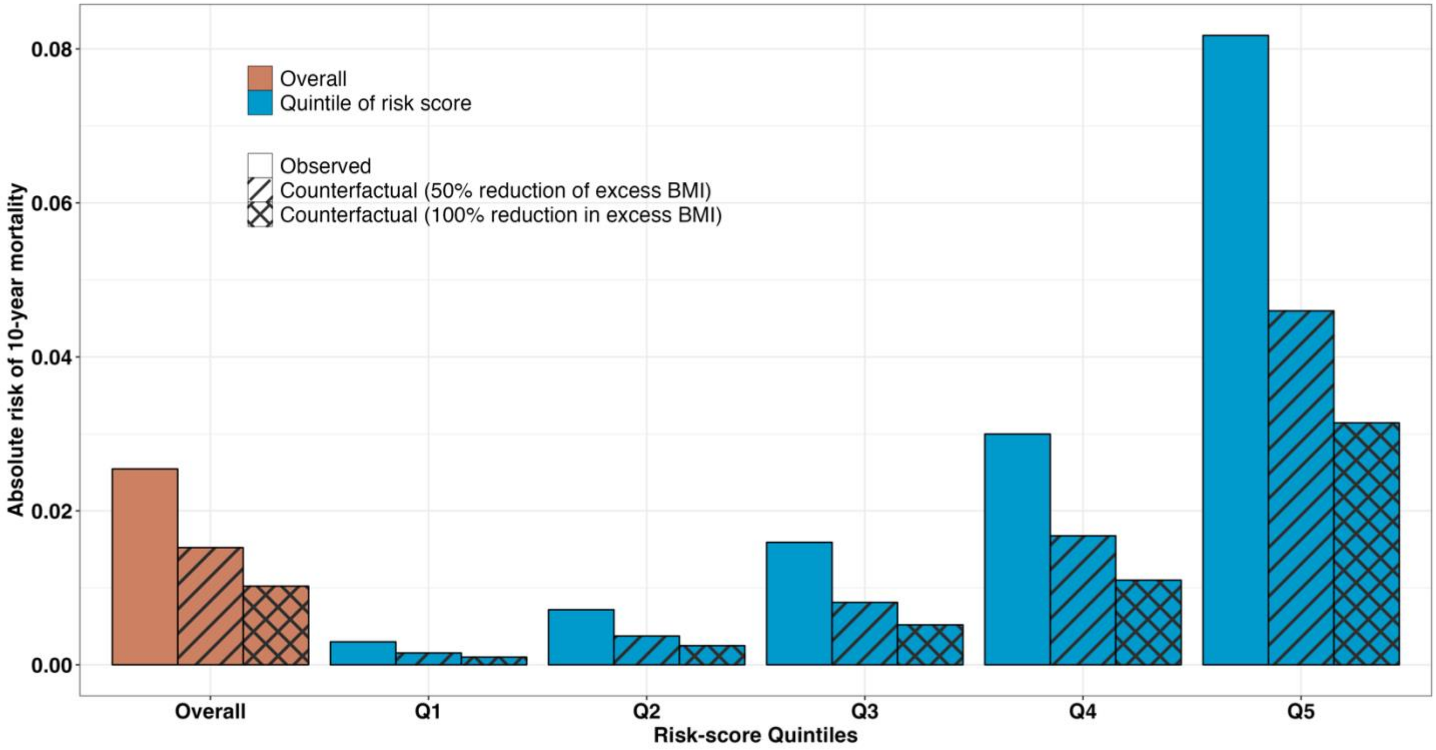

**Supplementary Figure 2: Observed and counterfactual absolute risks of 10-year mortality due to cancer for obese individuals in the non-Hispanic White population with an age range of 40-69 represented by the NHANES 1999-2006 Surveys**. Observed risks correspond to empirical proportions of deaths observed. Counterfactual risks correspond to average of the estimated absolute risks of the individuals associated with 50% and 100% reduction of excess BMI (compared to normal weight) where MR-derived estimates of relative-risks are used to represent underlying counterfactual effects. Results are shown for the overall population and stratified by the quintile of a risk score defined by other prominent risk factors of mortality. See Sections 1 and 4 of Supplemental Methods for details of the derivations.

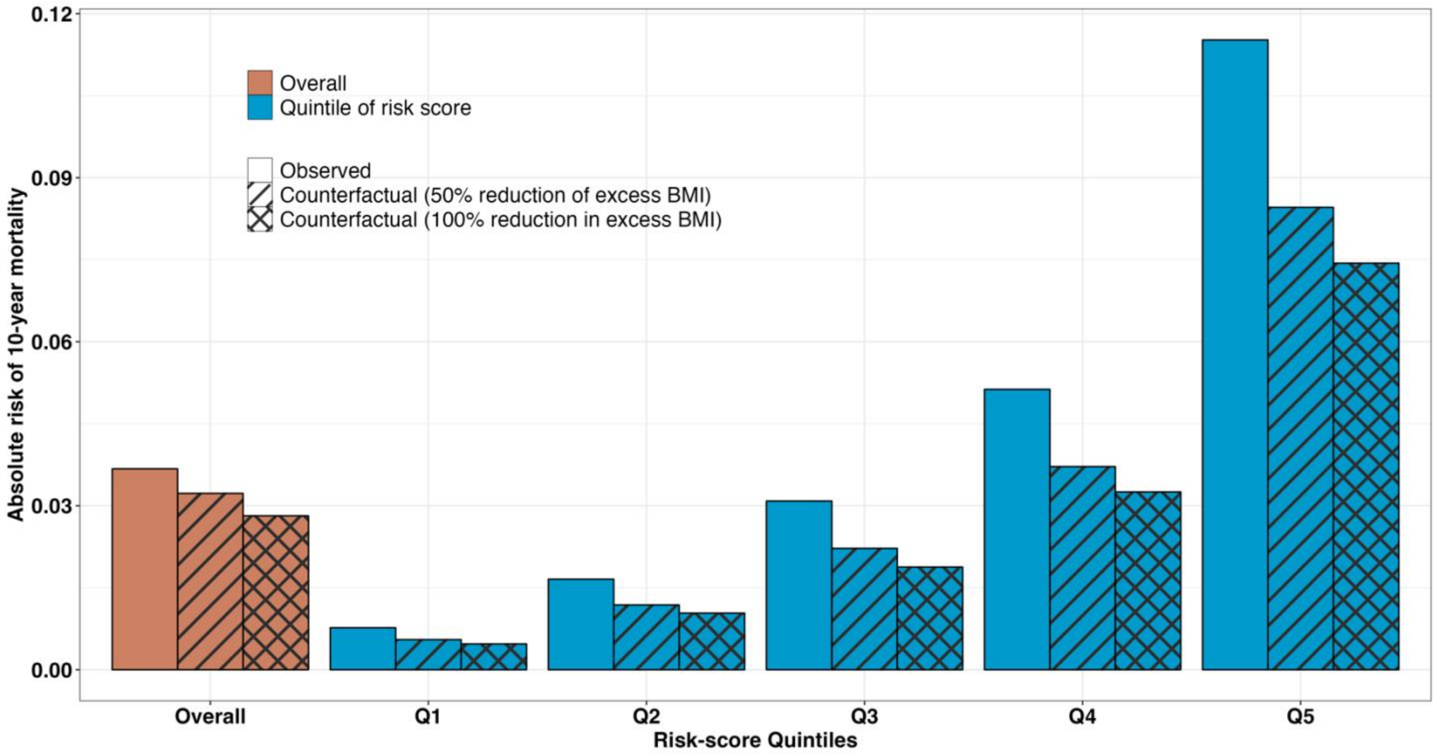

**Supplementary Figure 3: Observed and counterfactual absolute risks of 10-year all-cause mortality for obese individuals in the non-Hispanic White population with an age range of 40-69 represented by the NHANES 1999-2006 Surveys**. Observed risks correspond to empirical proportions of deaths observed. Counterfactual risks correspond to average of the estimated absolute risks of the individuals associated with 50% and 100% reduction of excess BMI (compared to normal weight) where estimates of <u>relative risks from pooled cohort analysis</u> are used to represent underlying counterfactual effects. Results are shown for the overall population and stratified by the quintile of a risk score defined by other prominent risk factors of mortality. See Sections 1 and 4 of Supplemental Methods for details of the derivations.

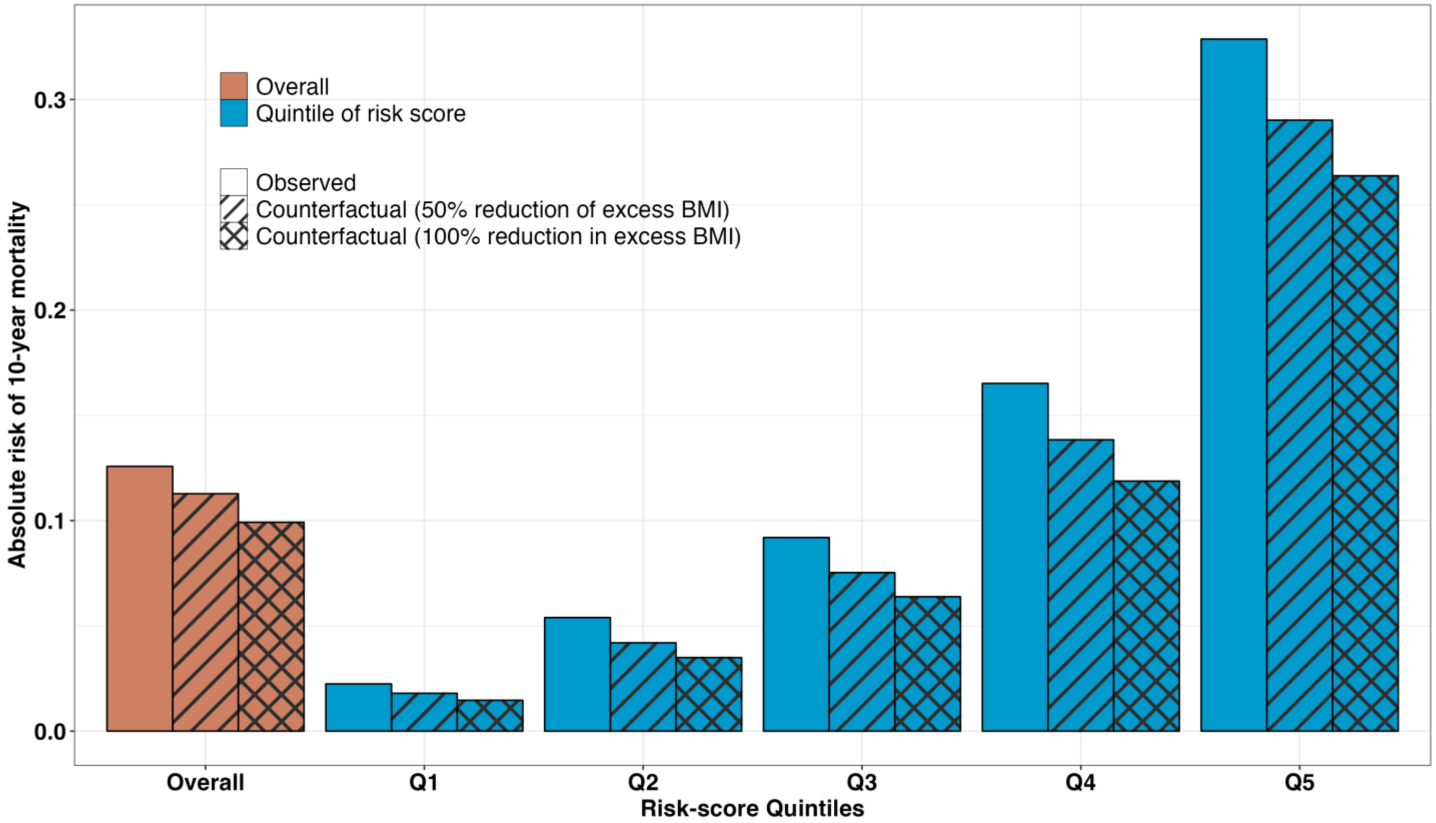

**Supplementary Figure 4: Observed and counterfactual absolute risks of 10-year mortality due to heart disease for obese individuals in the non-Hispanic White population with an age range of 40-69 represented by the NHANES 1999-2006 Surveys**. Observed risks correspond to empirical proportions of deaths observed. Counterfactual risks correspond to average of the estimated absolute risks of the individuals associated with 50% and 100% reduction of excess BMI (compared to normal weight) where estimates <u>of relative risks from pooled cohort analysis</u> are used to represent underlying counterfactual effects. Results are shown for the overall population and stratified by the quintile of a risk score defined by other prominent risk factors of mortality. See Sections 1 and 4 of Supplemental Methods for details of the derivations.

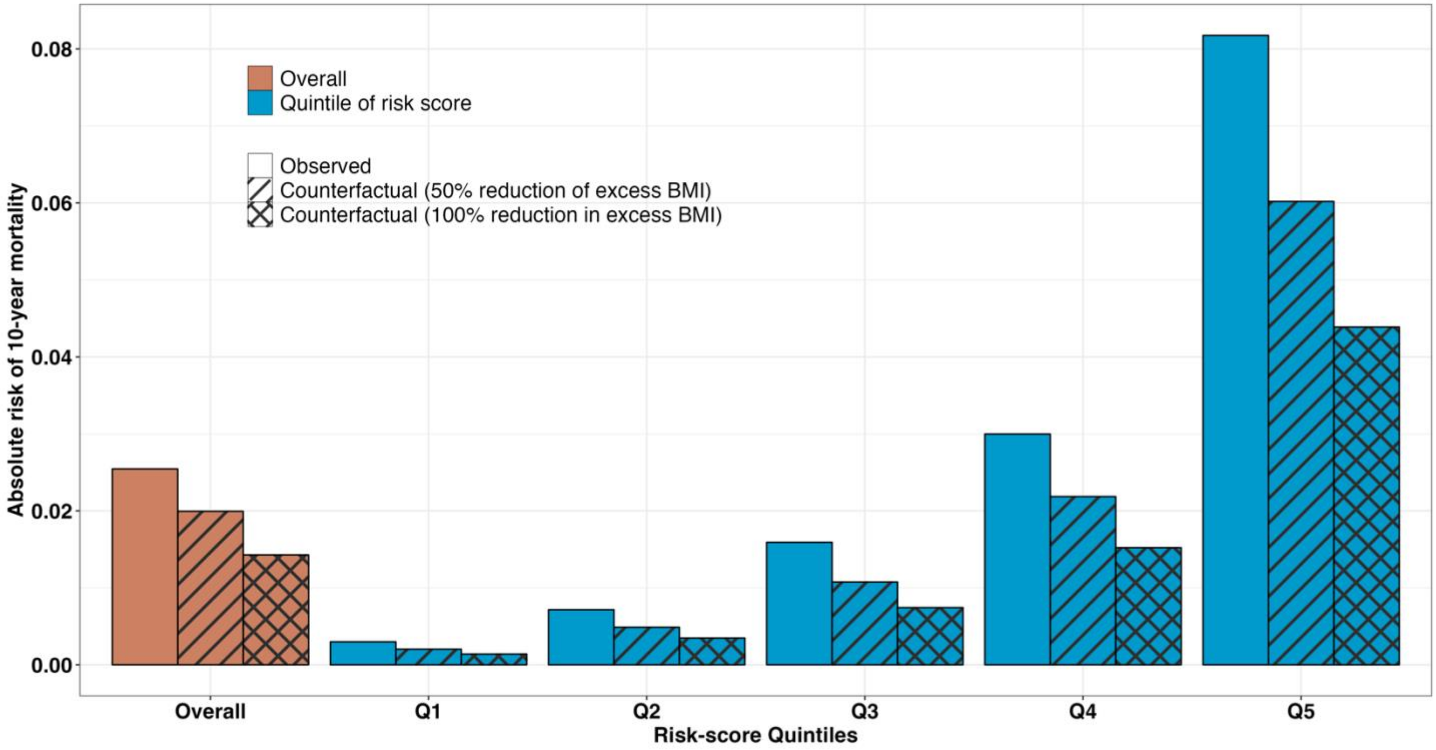

**Supplementary Figure 5: Observed and counterfactual absolute risks of 10-year mortality due to cancer for obese individuals in the non-Hispanic White population with an age range of 40-69 represented by the NHANES 1999-2006 Surveys**. Observed risks correspond to empirical proportions of deaths observed. Counterfactual risks correspond to average of the estimated absolute risks of the individuals associated with 50% and 100% reduction of excess BMI (compared to normal weight) where estimates of <u>relative risks from pooled cohort analysis</u> are used to represent underlying counterfactual effects. Results are shown for the overall population and stratified by the quintile of a risk score defined by other prominent risk factors of mortality. See Sections 1 and 4 of Supplemental Methods for details of the derivations.

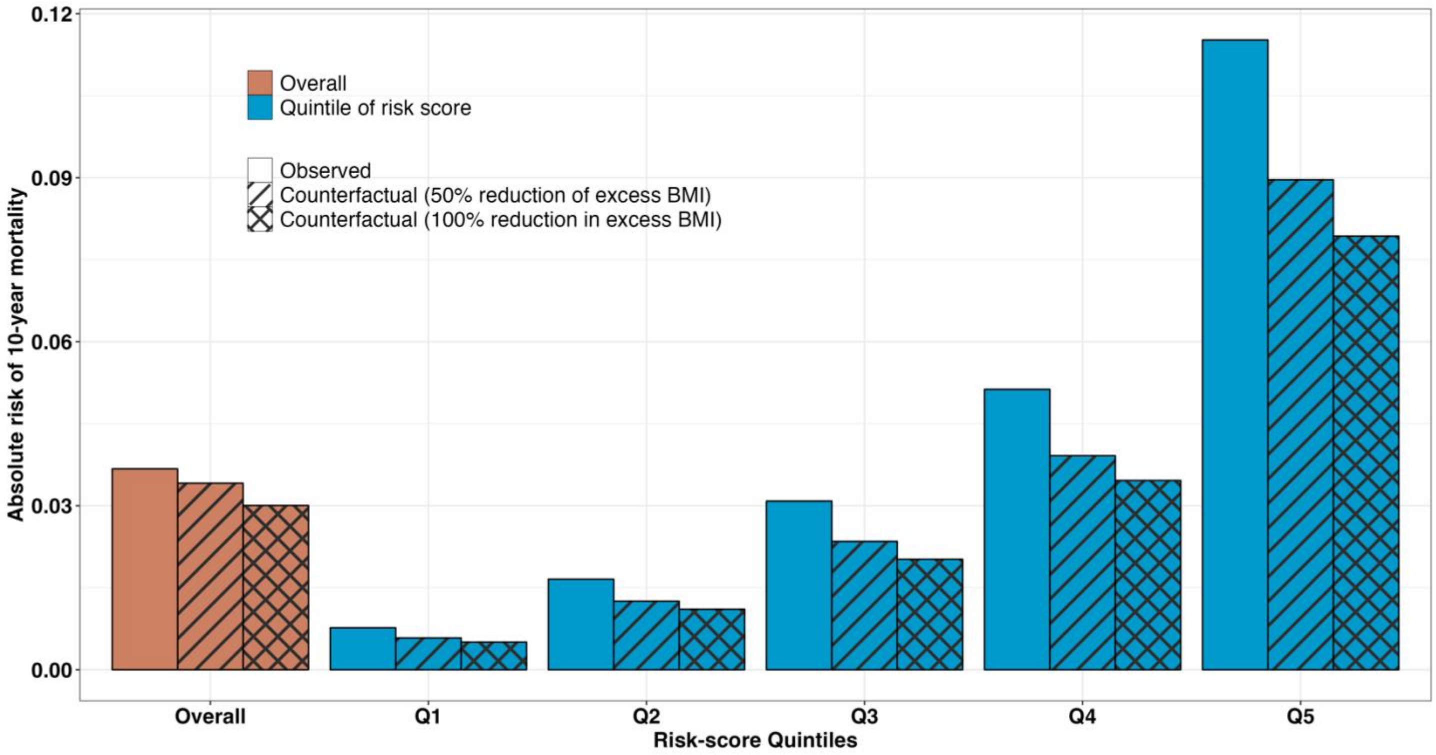
